# DEEP LEARNING BASED DETECTION OF URETHRAL STRICTURE: SEGMENTATION & CLASSIFICATION

**DOI:** 10.1101/2024.10.16.24315644

**Authors:** Nikhil Gurung, L Udaya Kumar, S N Chandrasekar, Sai Muthukumar V, Darshan Gera, Raghunatha Sharma, Ashwin Shekar P

## Abstract

**Purpose:** The retrograde urethrogram (RUG) has been a key diagnostic tool for over a century, remaining essential despite the availability of other imaging techniques for screening, diagnosis and follow up of Urethral strictures. However, interpretation of RUG images has to be done manually and needs experience on the part of the treating urologist, which calls for a common understanding of RUGs and presents a chance to improve stricture management in a practical way. Artificial intelligence (AI) algorithms present a novel way to prevent human discrepancy while concomitantly improving the accuracy of stricture identification and classification.

**Methods:** *Dataset:* We have used a balanced dataset which includes RUGs of 168 strictured cases and 178 non-strictured(healthy) cases.

*Task#1:* The primary requirement is to identify the Urethral region in any clinically obtained RUGs and detect the presence of stricture in it. We successfully deployed a Segmentation and Classification model to categorize the whole dataset as strictured or non-strictured RUGs.

*Task#2:* On obtaining superior accuracy, we effectively went on to identify the type of stricture based on their location, which is of clinical importance.

**Results:** With the above-mentioned available RUG dataset from 346 cases, we could train our Deep learning model and achieve a significant accuracy of 91.53% in detection and categorizing the type of stricture. At the end, a 10-fold cross-validation yielded an accuracy of about 86.66%.

**Conclusion:** Our attempts have successfully validated that using Deep learning (DL) tools, one could readily (i) Detect the presence of stricture in a given RUG and (ii) ultimately locate and classify these strictures effectively. Thus, these Deep learning tools could be of great clinical assistance for Urinary stricture related disease management.

## Introduction

Stricture in the male urethra is a common urological condition characterized by narrowing of the urethral passage that produces an obstruction to urine flow in men. Approximately 0.6% of males experience urethral stricture, and failure to treat it can result in serious urologic complications [1]. Globally, male urethral strictures are estimated to have a prevalence ranging from 229 to 627 per 100,000 individuals. In terms of healthcare utilization, male urethral strictures lead to around 5,000 hospital admissions and 1.5 million clinic visits annually in the US alone. In the United Kingdom, the prevalence is lower, estimated at 40 per 100,000 in men up to 65 years old and 100 per 100,000 in men over 65 years old [2]. The distribution of urethral stricture etiology varies across the world, and also with age [3]. The obstruction can enormously impair the patient’s quality of life and has a propensity to damage the entire urinary tract if left untreated It usually manifests as obstructive lower urinary tract symptoms (LUTS) like poor stream, straining to void and can sometimes lead to complications such as infection, bladder stones, fistulas, sepsis, and eventually renal failure.

Diagnostic investigations for suspected urethral strictures entail a series of procedures, such as cystoscopy, retrograde urethrography, and uroflowmetry. The retrograde urethrogram (RUG) has been a key diagnostic tool for over a century, remaining essential despite the availability of other imaging techniques and considered the current gold standard for evaluating strictures This method includes introducing a contrast dye into the urethra at the tip of the penis. The dye allows the doctor to see the whole anterior urethra and clearly define the strictured area. It is crucial to perform and interpret RUGs accurately for the correct diagnosis of urethral strictures and for planning surgical interventions [4]. The use of RUG in conjunction with other modalities can increase diagnostic accuracy in the evaluation of urethral stricture and aids in effective preoperative assessment and planning. However, interpretation of RUG is usually done manually and that can lead to a significant observer bias in identification and characterization of stricture.

In healthcare, Artificial Intelligence (AI) has emerged as a powerful tool for improving medical diagnostics, personalized treatment plans and patient outcomes. In particular, AI-based image analysis techniques have shown great promise in aiding medical professionals in the early detection and diagnosis of diseases, including cancer, cardiovascular issues and neurological disorders. AI tools are currently being used in the field of urology, especially in uro-oncology and urolithiasis with its role in reconstructive urology that includes urethral stricture still yet to be fully explored**[5-6]**. Herein, we propose a comprehensive AI tool in detecting and classifying these urethral Strictures from RUG.

## Materials & Methods

### DATA COLLECTION

Following approval from institutional REB, the retrospective RUG data was collected from the Department of Urology, Sri Sathya Sai Higher Medical Sciences (SSSIHMS), Prasanthigram, Andhra Pradesh. The dataset comprised 346 cases of patients treated at SSSIHMS for various urological health disorders and surgical procedures. We obtained a balanced dataset out of which **168 cases** were identified as strictured and **178 cases** were identified as non-strictured(healthy). The data was obtained in bitmap(bmp) format and anonymized using the *KerasOCR (Optical Character Recognition)* module which is used to detect and remove all patient identification text and ensure patient details confidentiality. The masks for training our Segmentation Model (UNet) were also generated using *ImageJ*. It is a popular, open-source image processing program designed for scientific and medical research which provides a wide range of image processing capabilities, such as filters, measurements, annotations, and transformations [7].

### TOOLS

#### Google Colab

It is a cloud-based platform offered by Google that enables users to develop, run, and share Python programs using a web browser. It features a Jupyter notebook interface and free access to processing resources such as GPU and TPU accelerators **[8]**. The dataset preparation and model training were performed in Colab. The accelerator used was the NVIDIA TESLA K80 12GB.

#### Frameworks

Keras and TensorFlow frameworks were used to build our deep learning models. Keras is an open-source, high-level neural networks API, written in Python and designed for simplicity and rapid prototyping **[9]**. It can run on top of other frameworks like TensorFlow, providing a user-friendly interface for building deep learning models. TensorFlow is an open-source machine learning framework that provides a comprehensive ecosystem for model development and deployment **[10]**. It supports a wide range of tasks, including deep learning and reinforcement learning, with tools for both high-level API access through Keras and low-level operations for custom algorithms. Optimized for performance on CPUs, GPUs, and TPUs, TensorFlow is widely used in the research community, providing the robust backend for Keras.

### Deep Learning Approach

**Figure 1** captures and renders an overview of the implementation of Deep learning technique at various stages in solving the current clinical problem. For training the Segmentation Model (**Task 1a)**, masks were created manually using ImageJ software so that only the region of interest, i.e., urethra is focused. Masks for all the cases were created for training the model. The data was then split into training, validation and testing sets in the following proportion: 70%, 20% and 10% respectively. Similarly, the dataset for the classification model (**Task 1b**) was prepared from the output masks from the segmentation model. So, a total of 346 data samples were used for optimization. Further, data augmentation with zoom, rotation and horizontal flip were applied to generalize the model. Similarly, **Task 2a & 2b** involving segmentation and classification model was developed for identifying the type of urethral Stricture.

**Figure 1:**
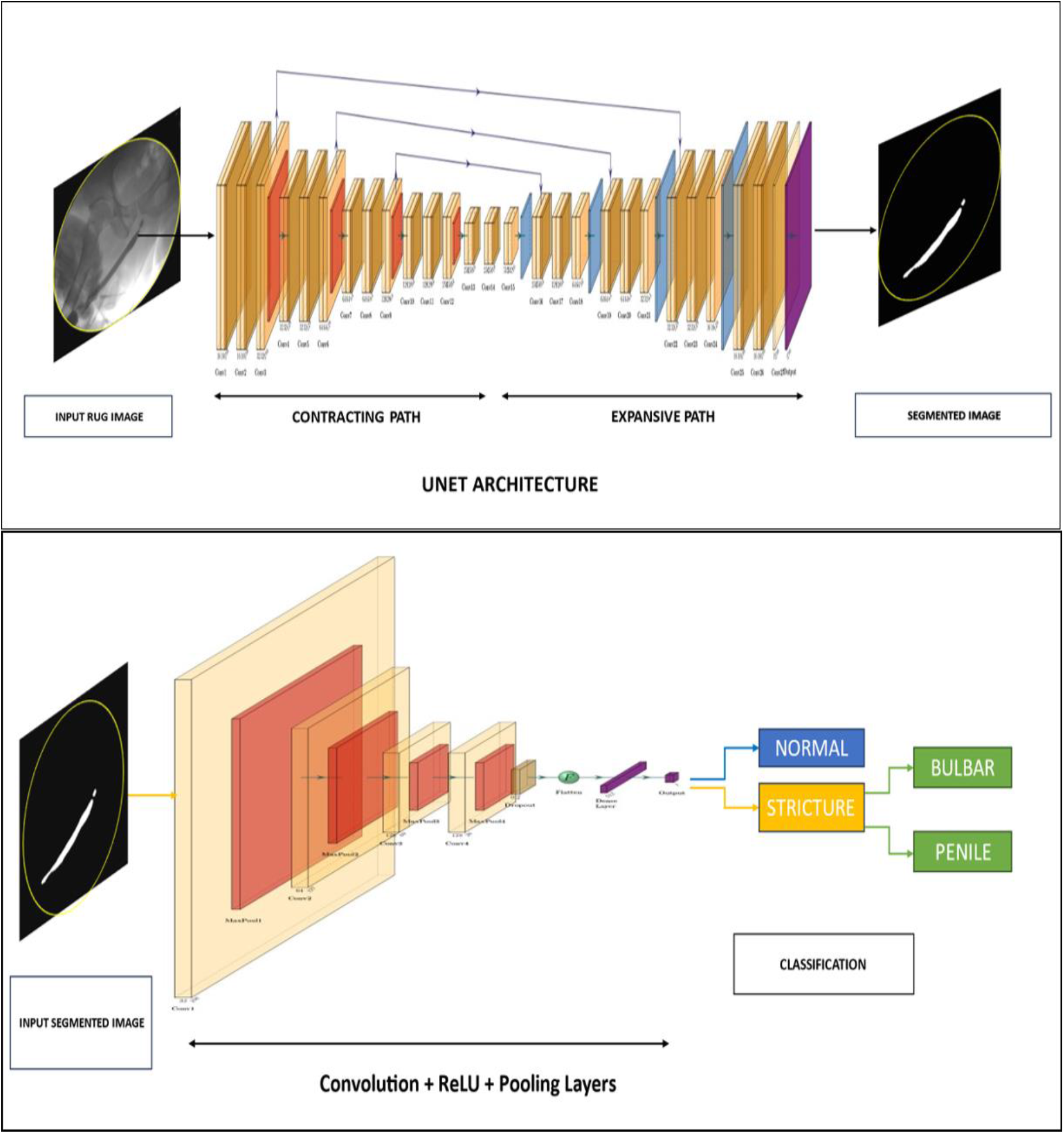
Overall model summary – an input RUG imge is fed to the UNET architecture which outputs the segmented image. The resultant image is then given as an input to the classification model which will eventually classifiy the given image as strictured or normal. Further the strictured image was sub classified into bulbar and penile stricture.

#### Segmentation model using UNet in *Task-1a & 2a*

UNet is a convolutional neural network used widely for the segmentation of biomedical images with encoding and decoding layers **[11]**. 2D convolutional layers with a kernel size of 3 was used. ReLU activation function was mainly used for hidden layers. The filter number increased from 16 and doubles up after every block in the contraction path (decoding block). The output of the present block was Max pooled to pass on to the next block. Dropout with the rate of 0.1-0.3 was used as a regularizer. The layer input from the decoding block was concatenated with the output of the Conv2D layers. The number of filters for the encoding blocks decreased from 128 to 16. The output of the Conv2D layer was taken with the sigmoid activation function with the kernel size of one.

#### Classification Model used in *Task-1b & 2b*

The model was built with four convolutional layers and four Max pooling layers. The first layer consists of 32 filters followed by 64 filters and the last two layers with 128 filters. After the last layer, a dropout layer was added to improve the performance. The model was trained with Adam optimizer with a learning rate of 0.001 and binary cross entropy was used as the loss function. ReLu activation function was used for hidden layers and the sigmoid function for the output layer. Various types of metrics accuracy, precision and recall along with the callbacks ‘ModelCheckpoint’ and ‘EarlyStopping’ were evaluated during the model training. Finally, a 10-fold Cross validation was also evaluated to check the behavior of the model.

## Results

### Task-1a: Segmentation Model performance

The U-net model was trained with a batch size of 32 along with the evaluation of accuracy, precision, recall, IoU (Intersection over Union) and dice coefficient metrics. The model achieved the highest dice coefficient value of *89*.*35%* by using various optimization techniques such as batch normalization and he-normalization. **Figure 2a** and **Figure 2b** showcases a representative output predicted by the segmentation model for the test dataset for both non-strictured and strictured RUG cases.

**Figure 2:**
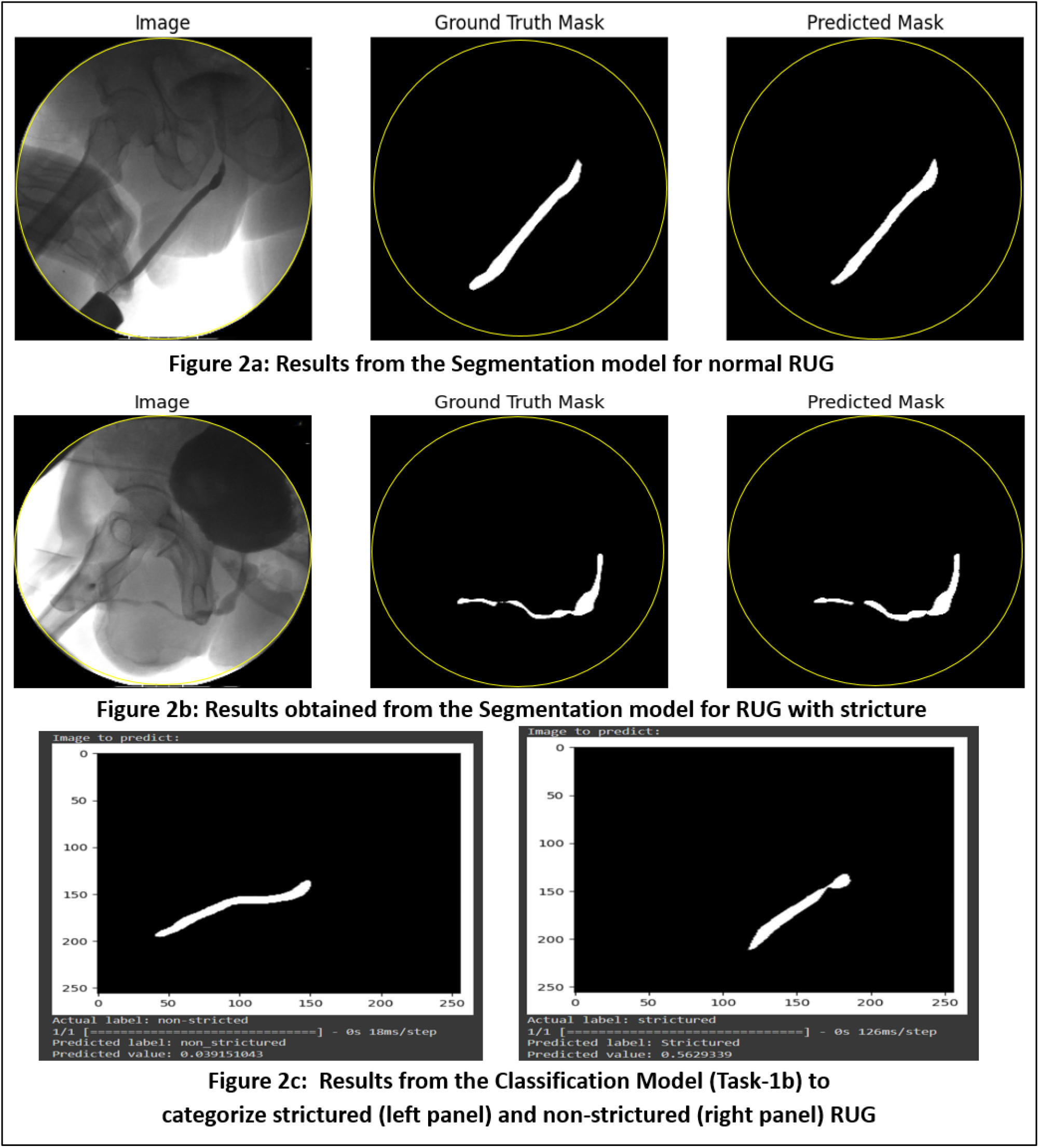
Summary of Task-1a & Task-1b which is the identification of Strictured and Non-Strictured Urethra in a given RUG data

### Task-1b: Classification Model performance

Following the Segmentation Model which could exclusively identify the urethral region from the complex urethrogram, our focus was to categorize RUG with and without stricture as shown in **Figure 2c**. Our classification model, trained for *30* epochs, achieved the highest accuracy value of *87*.*88%* with a binary cross entropy loss of *0*.*4082*. Finally, to avoid overfitting and increase the robustness of the model, we carried out the 10-fold cross validation which yielded a value of *82*.*48%*.

### *Task-2a & 2b:* Spatial Classification of Urethral Strictures

Classification of urethral strictures is based on two factors: location and its length. Among these, identifying the location of the stricture is a herculean task. In the current work, we have successfully attempted mapping the spatial information of the stricture. Typically, they are classified as (i) Penile strictures, (ii) Bulbar strictures and (iii) Penobulbar strictures **[12]**. Penile strictures spatially lies between the penoscrotal junction and the fossa navicularis. Bulbar strictures start at the penoscrotal junction to the bulbomembranous junction while Penobulbar strictures extend into the bulbar segment from the penile urethra, compromising lengthy urethral segments. With the available dataset we were able to classify Bulbar and Penile type stricture.

As shown in **figure 3**, the model could predict the stricture region as well as classify it into a particular type. This is a significant milestone achievement as the developed model can clearly and readily aid clinicians in pre-operative planning and disease prognostication.

**Figure 3:**
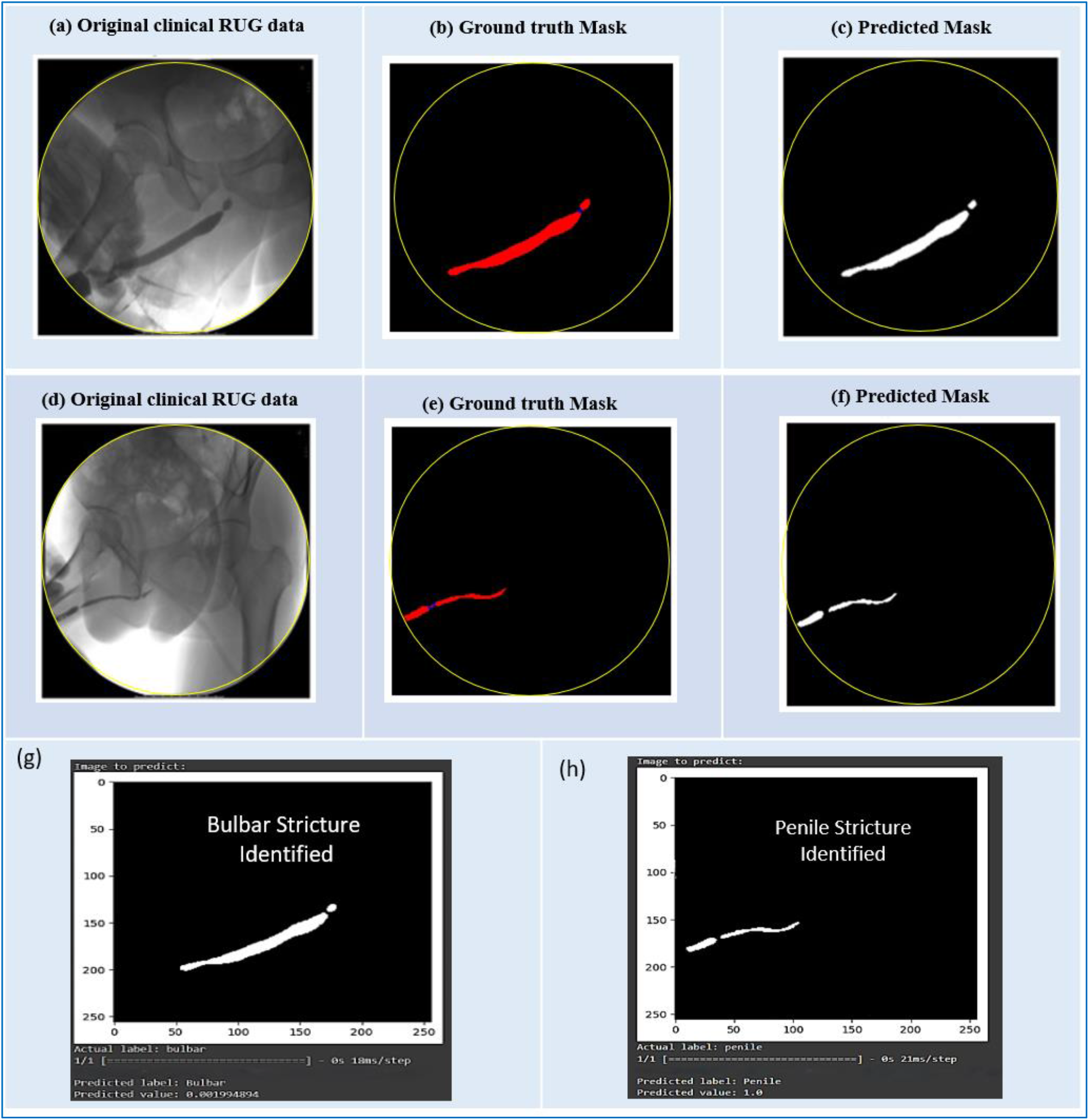
(a-f) shows the ground truth and predicted masks by the segmentation model for the bulbar and penile stricture and (g-h) shows the classification of two types of stricture.

The Deep Learning model for classifying the strictures were developed in similar lines as of Task-1a & 1b which yielded better accuracy. Technically, as shown in **Table 1**, the segmentation model yielded a dice coefficient value of *80*.*81%* and our classification model was able to achieve an accuracy of *91*.*53%*.

**TABLE I:**
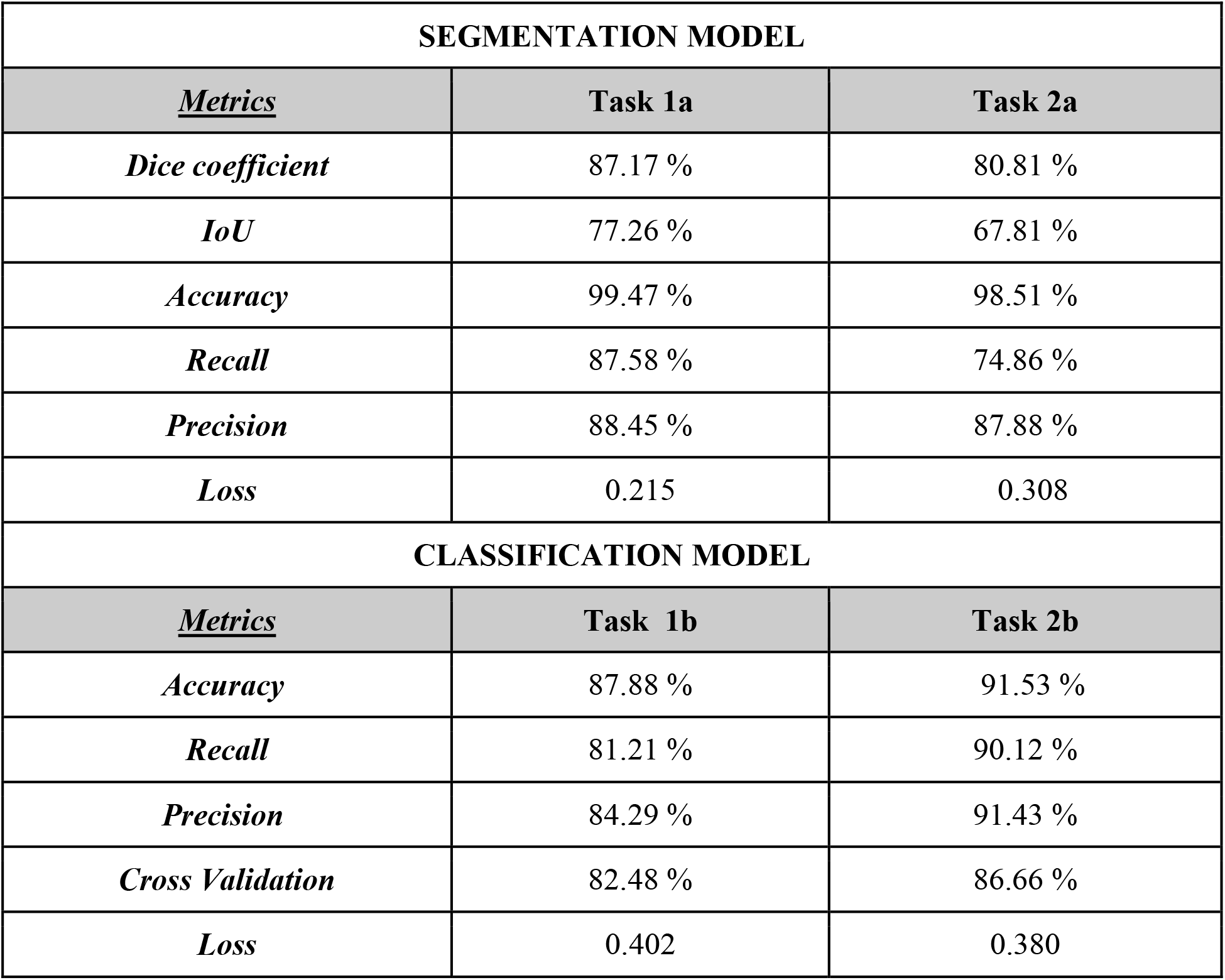
Collation of computational parameter obtained through deep learning process for the various tasks undertaken.

## Discussion

While machine learning based algorithms have been used extensively in a variety of sectors to categorize images in healthcare, relatively little work has been done in reconstructive urology, a field in which urethral stricture management plays a significant role. RUG is an invaluable test to evaluate and document the stricture and define stricture recurrence. Therefore, it is crucial to perform and interpret RUGs accurately for the correct diagnosis of urethral strictures and for planning surgical interventions. However, RUG has low sensitivity in determining the length, location, caliber, and other characteristics of strictures. It is generally suggested to be performed (or personally monitored) and interpreted by the treating urologist. Bach et al., study investigated the accuracy of retrograde urethrogram interpretation by primary physicians versus independent physicians [13]. The study aimed to evaluate the use of independent physician interpretations for pre-operative planning of urethral stricture surgery. The results indicated that primary physician-reported RUGs were more reliable than independently reported RUG. The interpretation of RUG images necessitates a greater level of expertise due to these limitations. As a result, it is crucial to foster a common understanding of RUGs, offering a valuable opportunity to enhance the practical management of strictures. There is potential for subjectivity in the interpretation of RUGs’ findings because they are frequently open to expert interpretation. Additionally, when physicians choose the course of treatment for urethral strictures based on the results of the RUG, there might be differences in how they explain the success rates of each procedure and the objectively preferred kind of reconstructive surgery to their patients. There is presently no “validated” method of evaluating an RUG, in contrast to certain other imaging categories that have distinct criteria for classification, including the CT classification of acute renal injury **[4]**. Because of this, interpreting an RUG requires professional judgment, which isn’t often available in healthcare settings.

In this report, we have detailed our implementation of AI tools for (i) Identification of Urethral stricture in a given clinical RUG data, (ii) detecting the site/region of the urethral stricture which was a very challenging task radiologically and (iii) ultimately, extended the AI model to do multi-label classification of stricture types. We could successfully execute this with the support of an independently in-house developed DL model which significantly offered more precision to this task. Using a balanced in-house dataset of 346 cases, we have achieved a detection accuracy of 91.53% and a 10-fold cross-validation accuracy of 86.66%. These results demonstrate the efficacy of deep learning models in identifying strictures and determining their type and location, enhancing clinical decision-making and reducing human error. Thus, it could be conceived as a decision supporting tool for the clinicians (irrespective of his/her experience) and help them identify the strictures efficiently. Leveraging our AI model, we can proficiently detect strictures, precisely determine their location and categorizing strictures based on its specific location. Our model is being developed further to ascertain the length of the stricture and offer prognostic insights for managing urethral strictures.

Earlier, a preliminary “proof of the concept” evaluation work was reported by Kim et al and their research group at Hospital for Sick Children, Division of Urology, Canada **[6]**. This was the first but very limited AI-based detection scheme reported for urethral strictures in 2022 using only 242 studies. They reported a very simple binary classification of mere presence or absence of strictures using a Convolutional neural network (CNN) model. Their work uses the whole RUG image for the classification, which may not give accurate results. Moreover, their dataset is not available in the public domain for utilizing and validating for other groups. Subsequently, we have found limited or no literature/reports by any other groups on these lines.

Unlike this deep learning model reported earlier by the Kim et al, our work is the first investigation to assess the possibility of using CNN-based algorithm in identifying RUGs and further classifying them based on location, which can provide valuable clinical information regarding strictures for further reconstruction. We aimed to develop a machine learning based algorithm to correctly identify the strictured area and categorize RUG images based on location as penile, bulbar and penobulbar. We demonstrated that we could achieve accuracy up to 87.88 % in classifying the stricture with normal RUG data and an accuracy of 91.53 % in classifying the bulbar and the penile stricture respectively. Our current model by segmenting the region of interest in the whole RUG where strictures are present and uses it further for classification of the stricture to a particular type is a comprehensive solution to the clinical requirement.

## Limitations

There are limits to this work, even if it is a novel attempt to identify and categorize urethral strictures according to their spatial location. The data was susceptible to possible overfitting from its training images because there weren’t many RUG images available for training. We have minimized this by performing data augmentation, which enables the model to function well with different types of altered images. While length is an important criterion during surgical planning a disease prognostication, there are other criteria also that will be need to be considered like length, etiology and luminal diameter. Nonetheless, we plan to further collaborate with other high-volume reconstructive urology centers and to prospectively obtain additional RUG images to develop more complex models that may characterize length and luminal diameter.

## Conclusion

Our study highlights the potential of AI-based deep learning tools in improving the accuracy and reliability of Retrograde UrethroGram (RUG) interpretations for detecting and classifying urethral strictures. Integrating these tools into pre-operative evaluations could significantly improve urethral stricture management. This advancement promises better patient outcomes and streamlined clinical workflows. Future work will focus on extending the model to accurately identify stricture length, further enhancing its clinical utility.

## Data Availability

All data produced in the present study are available upon reasonable request to the authors

## Acknowledgements

SSSIHL: Authors at SSSIHL acknowledge inspiration driven by our Founder Chancellor-Sri Bhagawan Sathya Sai Baba Varu and SSSCT Trustees for building a research infrastructure and ecosystem with their generous support and funding.

## Research involving human participants

No animals or human participants were required for this study.

## Informed Consent

Informed consent was waived as this study is a retrospective analysis of imaging.

